# The potential for bike riding across entire cities: quantifying spatial variation in interest in bike riding

**DOI:** 10.1101/2021.03.14.21253340

**Authors:** Lauren K Pearson, Joanna Dipnall, Belinda Gabbe, Sandy Braaf, Shelley White, Melissa Backhouse, Ben Beck

## Abstract

**Background:** Riding a bike is beneficial for health, the environment and for reducing traffic congestion. Despite this, bike riding participation in the state of Victoria, Australia, is low. To inform planning and practice, there is a need to understand the proportion of the population (the ‘near-market’) that are interested in riding a bike, and how this varies across regions. The Geller typology classifies individuals into one of four groups, based on their confidence to ride a bike in various infrastructure types, and frequency of bike riding. The typology has been used at a city, state and country-wide scale, however not at a smaller spatial scale. We aimed to characterise and quantify the distribution of the Geller typology within Local Government Areas (LGAs) in the state of Victoria, Australia.

**Methods:** An online survey was conducted in 37 LGAs in Victoria, including all LGAs in Greater Melbourne, and a selection of six key regional centres. Participants were recruited from an opt-in online research company panel with the objective of recruiting a representative sample of adults across each LGA. The Geller typology classified individuals as either: ‘Strong and Fearless, ‘Enthused and Confident’, ‘Interested but Concerned’, or ‘No Way No How’. ‘Interested but Concerned’ participants are those that would ride a bike if protected infrastructure were provided.

**Results:** The survey was completed by 3999 individuals. Most participants owned a bike (58%), however only 20% rode at least once per week. The distribution of the Geller groups was: ‘Strong and Fearless’ (3%), ‘Enthused and Confident’ (3%), ‘Interested but Concerned’ (78%), and ‘No Way No How’ (16%). While variation in the distributions of the Geller groups was observed between LGAs within Greater Melbourne., the ‘Interested but Concerned’ group, reflecting people who are comfortable riding only in protected lanes or off-road paths, was high across all LGAs and all demographic sub-groups. Even though the frequency of riding a bike was lower in women, interest in riding a bike was high and comparable to men. Participants who resided in the outer urban fringe regions of Greater Melbourne had high interest, but low participation in bike riding.

**Conclusions:** While there was variation in interest in bike riding across an entire metropolitan region and across population groups, interest was high across all areas and demographics. Our results show the potential for substantial increases in cycling participation, but only when high-quality cycling infrastructure is provided. Further research is required to understand the policy and practice barriers to equitable provision of protected infrastructure.

## INTRODUCTION

Increasing participation in bike riding is well established as being beneficial for public health, the environment and traffic congestion (1-5). The majority of populated areas of Australia exhibit ideal conditions for riding a bicycle, including relatively flat topography and in most areas, a mild climate. Despite this, participation in bike riding remains low in Australia compared to other international settings (6). To increase bike riding participation and to ensure that planning and practice efforts are targeted to the whole community, there is a need to identify specific sub-groups of the population that can be classified as the bike riding ‘near-market’; individuals who would like to start riding a bike. Bike rider typologies are commonly used to achieve this, by segmenting populations into distinct groups with shared characteristics.

The most commonly used bicyclist typology is the ‘Four Types of Cyclist’ typology, first introduced in 2006 by Roger Geller (7) and later refined by Dill & McNeil (8). This classifies people into one of four groups based on their confidence to ride a bike in various infrastructure types, their interest in riding a bike and if they had ridden a bike in the past month. One of the four categories are people who are ‘Interested but Concerned’. Interested but Concerned participants are those that would ride a bike if protected infrastructure were provided. The typology has been used to quantify potentially latent groups of bicyclists in cities, states, and countries. While the use of typologies to quantify the near-market of bicyclists across large cities and regions has been helpful in informing bike riding strategies, prior studies have not explored whether interest in bike riding varies within these geographies, such as across smaller spatial areas of cities and states (8-11). The availability and quality of transport infrastructure often varies within cities depending on zoning, distance from the city, topography, socioeconomic status and community demographics (12-14). Identifying interest in bike riding at this small spatial scale is useful in understanding geographical variation in bike riding potential and how protected infrastructure can be implemented to maximise bike riding participation and address inequities in participation and infrastructure. To our knowledge, previous literature has not quantified the Geller typology groups at a spatial scale smaller than cities, and has not been applied in Australia.

In this study, we aimed to quantify and identify the characteristics of the ‘Four Types of Cyclist’ in all Local Government Areas (LGAs) within Greater Melbourne, and a selection of regional centres in the state of Victoria, Australia.

## METHODS

### Study design

We conducted a cross-sectional online survey in the state of Victoria, Australia. The objective was to recruit a representative sample of adults (aged 18 years and older) across each local government area. Data collection occurred over the period of August 12^th^ to September 10^th^, 2020. Data collection occurred during a period of restrictions due to the COVID-19 pandemic. Many Victorian workplaces were closed and physical distancing was enforced (15), but there were no restrictions placed on leaving the home during this period.

In line with values-based messaging for health promotion (16), we adopted the term “bike riding” rather than “cycling", and their equivalents, in this study to ensure inclusivity and avoid association with competitive cycling (17).

### Setting

Victoria has a population of 6.7 million (18) of which 67% reside in the Greater Melbourne area (19). As of 2018, bike riding comprised 3% of all weekday trips in metropolitan Melbourne (20). LGAs were chosen as the geographical area for analysis. LGAs in Australia are defined subdivisions of states that are under the jurisdiction of a particular Local Government (21). In Victoria, 85% of the road network is made up of local roads, which are maintained by Local Governments (22). For this reason, Local Governments play an essential role in the planning and maintenance of bicycling infrastructure. Data were collected in each LGA within Greater Melbourne (n=31) and select regional LGAs (n=6) (Figure 1).

**Figure 1.**
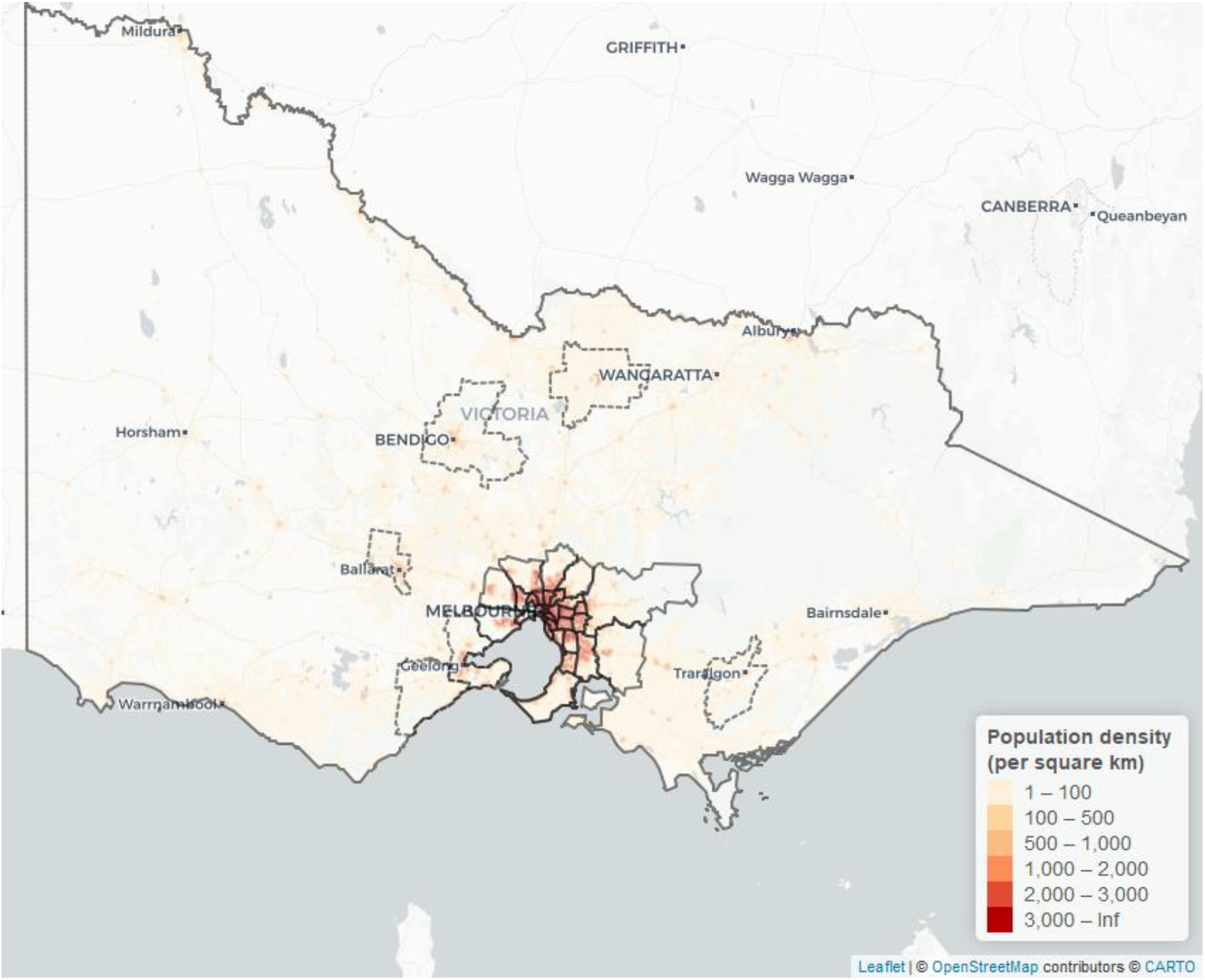
Map of included LGAs and population density within the state of Victoria (Regional LGAs = dotted border, Greater Melbourne LGAs = solid border)

Greater Melbourne LGAs were chosen specifically due to the high density of population (see Figure 1, Figure 2), and potential to improve bike riding participation rates and infrastructure. Similarly, data were collected in six regional centres with higher populations, higher population density and relatively flat topography. These areas also had the potential for increased participation in bike riding. Achieving a representative sample in rural/regional LGAs other than those selected was not feasible due to small survey panel sizes in these regions.

**Figure 2.**
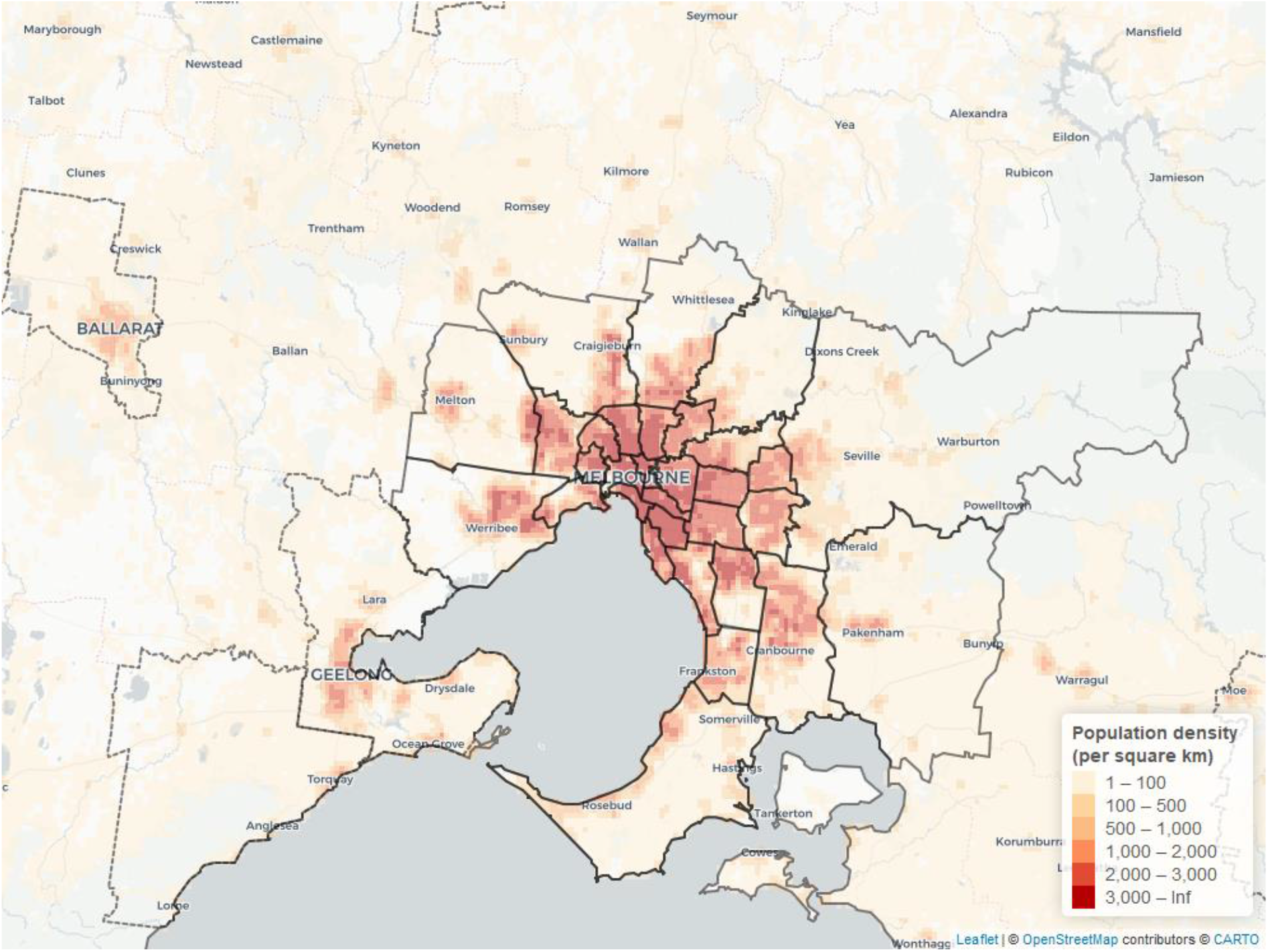
Map of population density in Greater Melbourne (Greater Melbourne LGAs = solid line, Regional LGAs = dotted line)

### Survey design

A cross-sectional online survey was developed by the study authors, and administered through Qualtrics Survey Software (23). The full survey is provided as Supplementary Material A. The survey included questions about demographics, bike riding frequency and behaviours, and a set of questions to categorise participants into one of the four Geller cycling topologies (7, 8). The Geller typology uses descriptions and illustrations of a variety of street environments, levels of bike riding infrastructure and traffic speeds and asks participants how comfortable they would be riding a bike in those conditions on a 4-point Likert scale, from 1 = “Very uncomfortable” to 4 = “Very comfortable". Participants are also asked their level of agreement with the statement “I would like to cycle more", with a 4-point Likert scale from 1 = “Strongly disagree” to 4 = “Strongly agree". The answers to these scenarios distinguish which of the four bike riding typologies the participant belongs to. Groups consist of people who are “Strong and Fearless", “Enthused and Confident, “Interested but Concerned” or “No Way No How". Strong and Fearless participants are comfortable riding a bike on any infrastructure, including on roads with no protected bicycling infrastructure. Enthused and Confident participants are comfortable on non-residential streets with painted on-road bike lanes (separating bikes from traffic without protection). Interested but Concerned participants are comfortable riding a bike on protected bicycling infrastructure only, while No Way No How participants are not comfortable riding a bike in any setting.

Adaptations were made to the Geller Typology for use in Australia. These included the conversion of miles per hour to kilometres per hour, and change of the term “striped bike lane” to “painted bike lane". Each question included a figure to allow the participant to better understand the scenario and context (see Figure 3). Photos were not used as to avoid associations with particular areas, or other factors that could influence bike riding comfort. The Geller typology was chosen for this study for its ability to inform policy and practice, and to allow for comparisons to studies in other settings that have used the same questions. Questions regarding frequency of bike riding and trip purpose were asked from the perspective of before COVID-19 restrictions were in place (see Supplementary Material).

**Figure 3.**
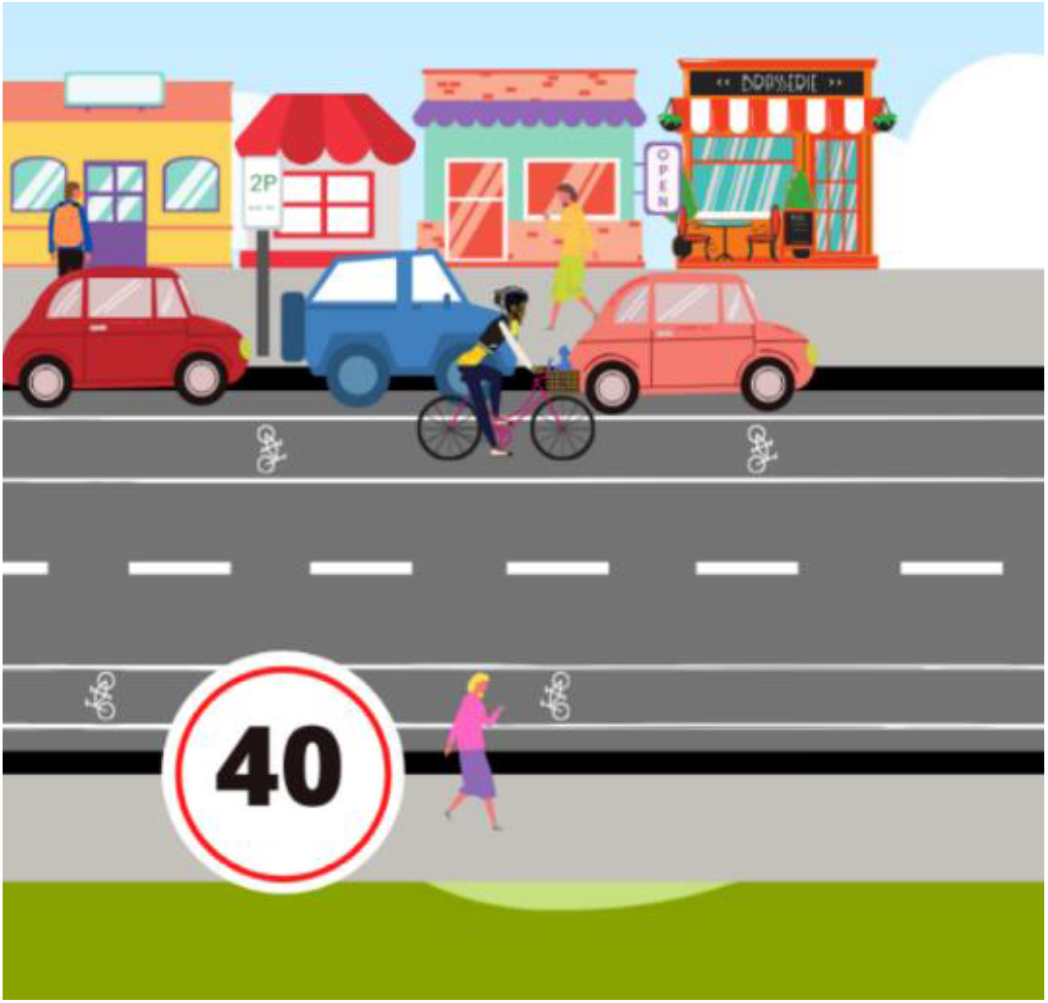
Example image alongside Geller Typology questions - Question 3: “How comfortable would you feel riding a bike on a two-lane commercial shopping street with traffic speeds of 30-40 km/h, on-street parking and a painted bike-lane?” Image developed by the authors.

Demographic data included age category (18-24 years, 25-34 years, 35-44 years, 45-64 years, 65-74 years, 75 and above years), gender (female, male, non-binary, other (contents specified by participant)), individual gross annual income from 2019 in $AUD ($1-$10,399, $10,400-$20,799, $20,800-$31,199, $31,200-$41,599, $41,600-$51,999, $52,000-$64,999, $65,000-$77,999, $78,000-$103,999 and $104,000 or more), whether the person spoke a language other than English at home, and Victorian postcode of residence. Postcodes do not form part of the Australian Statistical Geography Standard (24) and do not necessarily uniquely fit within a single LGA. Therefore, where a participant resided in a postcode bounded across multiple LGAS (*n*=79 postcodes), they were directed to a map of their postcode with LGAs boundaries and asked to select the LGA they resided in.

### Sampling and recruitment

We recruited people 18 years of age or older who resided in one of the LGAs in Greater Melbourne or one of the six regional LGAs. The aim was to achieve a representative sample size within each LGA, so that typologies between LGAs could be compared. Sample size was calculated for each of the LGAs based on the 2016 Australian Bureau of Statistics Census 2016 data (25) and using a 95% confidence level and a 2% margin of error, totalling *n* = 3351. The survey was distributed online via email to a sample obtained from a market research company, The Online Research Unit (ORU). The ORU panels are built using a variety of recruitment methods to ensure adequate general population representation and reduce selection bias: through post, phone, print and online. Invitations were sent in batches from Monday to Friday, with more invitations being sent on Friday to take account of the longer time period over the weekend where no email invitations were sent. By sending the invitations in batches, this reduced bias towards early responders and minimised response bias by distributing responses over different days. Participants who had partially responded to the survey or not entered the survey were sent a maximum of two reminders over the data collection period to reduce non-response bias. In order to monitor the Victorian population sample representativeness, the ORU were sent daily reports for the age and gender distributions for the completed survey sample to date. Where particular groups were lower than the population, ‘boosting’ was performed by sending further invitations and reminders those demographics not meeting the required proportions for sample representativeness. Age and gender representation at the LGA level was not possible to monitor due to restrictions on information sharing restrictions between the ORU and Monash University. Oversampling was performed for small LGA sub-populations to allow for more reliable estimates and to avoid sampling bias due to under-coverage. When sample sizes were achieved, no further invitations were sent to participants that resided in these areas.

### Statistical analysis

Responses were checked for duplicates, of which none were found. Completion times were checked for outliers to identify any responses completed in a time that was not feasible, and removed if so. Further cleaning of potentially straight-lined responses was not possible as these responses may have been valid.

Results for the overall sample were weighted to reflect the correct population representation of each LGA to account for oversampling in any of the LGAs and to remove any sampling bias. Statistical analysis was performed in the statistical program R and the integrated development environment RStudio (26) using the *survey* library for analysis of weighted data (27). Survey demographic data were compared to the aggregate population data for each of the included LGAs from the 2016 Census (25), with the exception of data relating to speaking a language other than English at home. This was not available for the selected LGA populations, and instead was obtained from a separate report and based on the whole of the Victorian population (28). Maps of population density (Figure 1 and Figure 2) were produced based on Australian Bureau of Statistics population density data (29), using the *leaflet* package in R (26, 30).Categorisation of participants into Geller Typology groups was determined by responses to a selection of survey questions, as described by Dill & McNeil (8). This process is described in Supplementary Materials E. Descriptive statistics and frequency tables of weighted data were used to describe the bike riding behaviours and frequency of each Geller Typology, and to make comparisons between age, gender and region of residence. Chi-square tests were used to examine differences in the distribution of the Geller Typology within age, income, and gender groups, and between Greater Melbourne and regional Victoria.

### Ethics

The project was approved by the (*NAME REMOVED FOR PEER-REVIEW*) Human Research Ethics Committee (Project ID: 25346).

## RESULTS

In total, 41,000 invitations and 20,000 reminders were sent by the ORU, with a response rate of 10%. Of the 4067 participants who started the survey, 3999 (98%) completed all sections and were included in the final analysis. The age and gender of the weighted sample were generally consistent with the aggregated population of the LGAs included in the study, with only the 65-74-year age group being 6.9% higher in the sample than the target population (see Supplementary Materials C). The weighted sample had a higher income than the target population, with 15% of participants earning over $104,000 per year compared to 7.2% of the target population. In the sample, 12% spoke a language other than English at home, compared to 26% in Victoria (see Supplementary Materials C).

Over half of all participants owned a bike (57%), however only 20% rode their bike at least once per week. Of participants that rode their bike at least once a year, most rode for recreation only (72%). Over three quarters of the sample (78%) were classified as Interested but Concerned, 2.8% as Strong and Fearless, 3.2% as Enthused and Confident and 16% as No Way No How (see Figure 4). The Interested but Concerned group comprised a higher proportion of women (48%) than the Strong and Fearless (28%) and Enthused and Confident groups (25%) (Table 1). Over half of this group owned a bike (63%), and most rode for recreation only (73%), and rode 1-3 times per year (31%). Twenty-six percent of participants in this group had not ridden a bike in the past 12 months, showing a potentially latent group of bicyclists.

**Table 1.** Geller Group characteristics based on sample weighted to the Victorian population (25)

|  | Strong and Fearless | Enthused and Confident | Interested but Concerned | No Way No How | Total Weighted Sample |
| --- | --- | --- | --- | --- | --- |
| **Proportion of weighted sample** | 2.8% | 3.2% | 78.2% | 15.7% | 100% |
| **Gender\*** |  |  |  |  |  |
| Female | 28.0% | 23.6% | 48.3% | 59.8% | 49.1% |
| Male | 71.1% | 74.8% | 51.3% | 39.6% | 50.5% |
| Non-binary | 0.9% | 0.8% | 0.3% | 0.3% | 0.3% |
| Other | 0.0% | 0.0% | 0.0% | 0.0% | 0.0% |
| Prefer not to say | 0.0% | 0.8% | 0.1% | 0.3% | 0.1% |
| **Age category** |  |  |  |  |  |
| 18 – 34 years | 31.7% | 38.0% | 31.8% | 17.7% | 33.0% |
| 35 – 54 years | 37.3% | 34.3% | 33.1% | 20.3% | 34.6% |
| 55 – 74 years | 30.1% | 24.5% | 32.2% | 50.6% | 24.0% |
| 75+ years | 0.9% | 3.2% | 2.9% | 11.4% | 8.4% |
| **Annual income** |  |  |  |  |  |
| \$1 - \$20,799 | 11.1% | 15.1% | 16.2% | 22.4% | 32.6% |
| \$20,800 - \$41,599 | 20.4% | 14.0% | 19.1% | 30.3% | 25.7% |
| \$41,600 - \$64,999 | 16.8% | 30.6% | 22.8% | 16.5% | 20.1% |
| \$65,000 - \$103,999 | 29.6% | 21.2% | 26.8% | 21.0% | 14.3% |
| \$104,000 + | 22.1% | 19.1% | 15.1% | 9.9% | 7.4% |
| **Bike ownership** |  |  |  |  |  |
| Yes | 70.6% | 79.9% | 63.4% | 20.4% | 57.3% |
| No | 29.4% | 20.1% | 36.6% | 79.6% | 42.7% |
| **Frequency of bike riding** |  |  |  |  |  |
| Never | 20.1% | 8.5% | 25.6% | 76.1% | 32.8% |
| 1 - 3 times per year | 15.2% | 23.1% | 30.5% | 12.7% | 27.1% |
| 1 - 3 times per month | 15.5% | 21.9% | 23.1% | 5.9% | 20.1% |
| 1 - 3 times per week | 25.0% | 24.0% | 15.7% | 4.3% | 14.5% |
| 4 + days per week | 24.2% | 22.4% | 5.0% | 0.9% | 5.5% |
| **Primary trip purpose\*\*** |  |  |  |  |  |
| Transport | 11.9% | 7.2% | 6.5% | 16.1% | 7.3% |
| Recreation | 48.7% | 51.7% | 73.4% | 72.9% | 71.6% |
| Both | 39.4% | 41.1% | 20.1% | 11.0% | 21.1% |

\*For the counts of participants who identified as “non-binary” or “other”, or reported “prefer not to say”, robust analysis could not be performed due to small sample sizes
\*\*based on participants who had ridden a bike in the 12 months previous to COVID-19 restrictions

**Figure 4.**
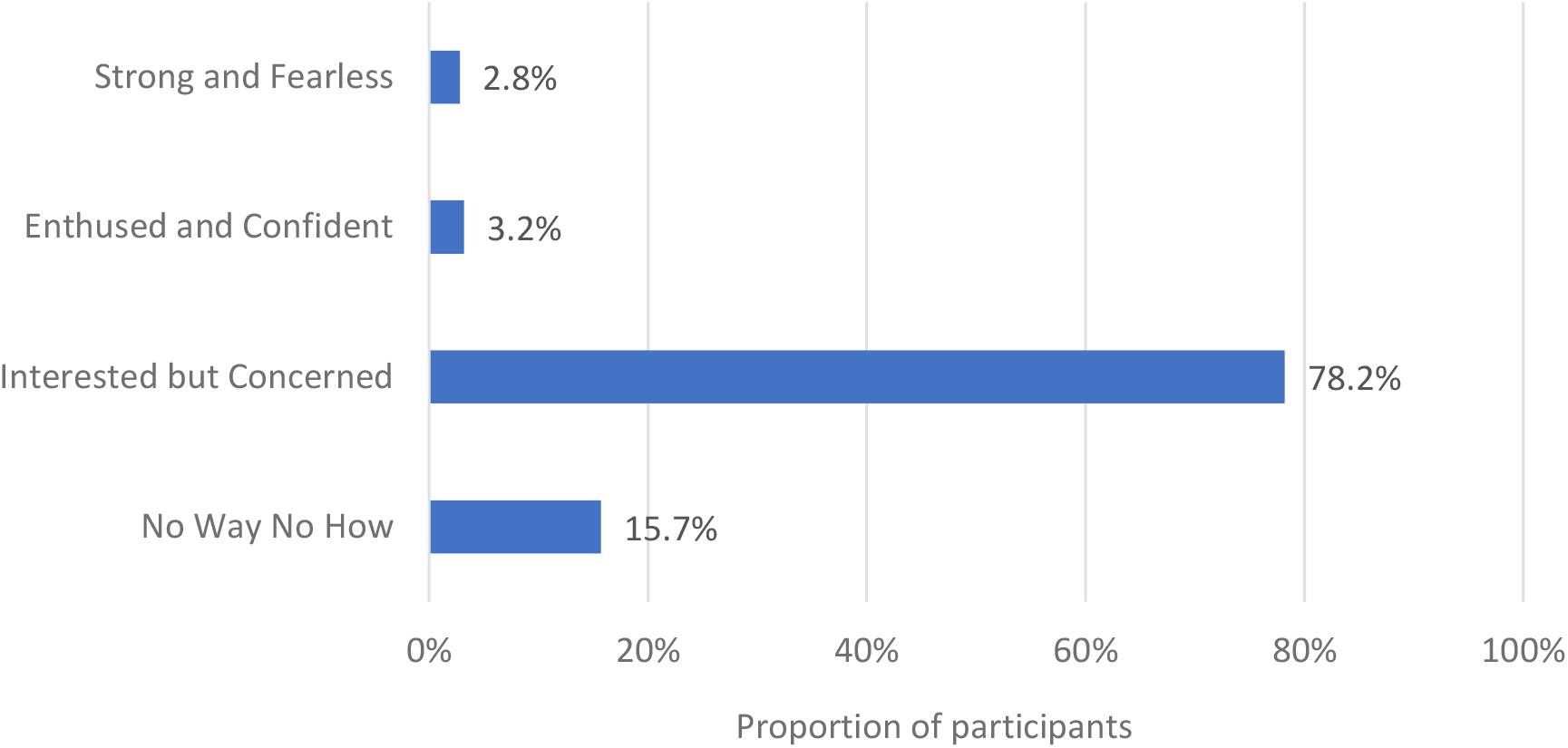
Bar graph of the distribution of Geller groups across weighted study sample (n = 3999)

### Within Greater Melbourne comparisons

In Greater Melbourne, the Interested but Concerned group was consistently the largest group across all LGAs, ranging from 66% to 88% (Figure 5). Many of the LGAs on the outer-fringe area of Greater Melbourne had higher proportions of Interested but Concerned participants. The proportion of Strong and Fearless were low in all LGAs, ranging from 0.0 to 7.1%. Similarly, proportions of Enthused and Confident participants were low, ranging from 0.0% to 7.3%. Proportions of No Way No How participants ranged from 2.7% to 23%.

**Figure 5.**
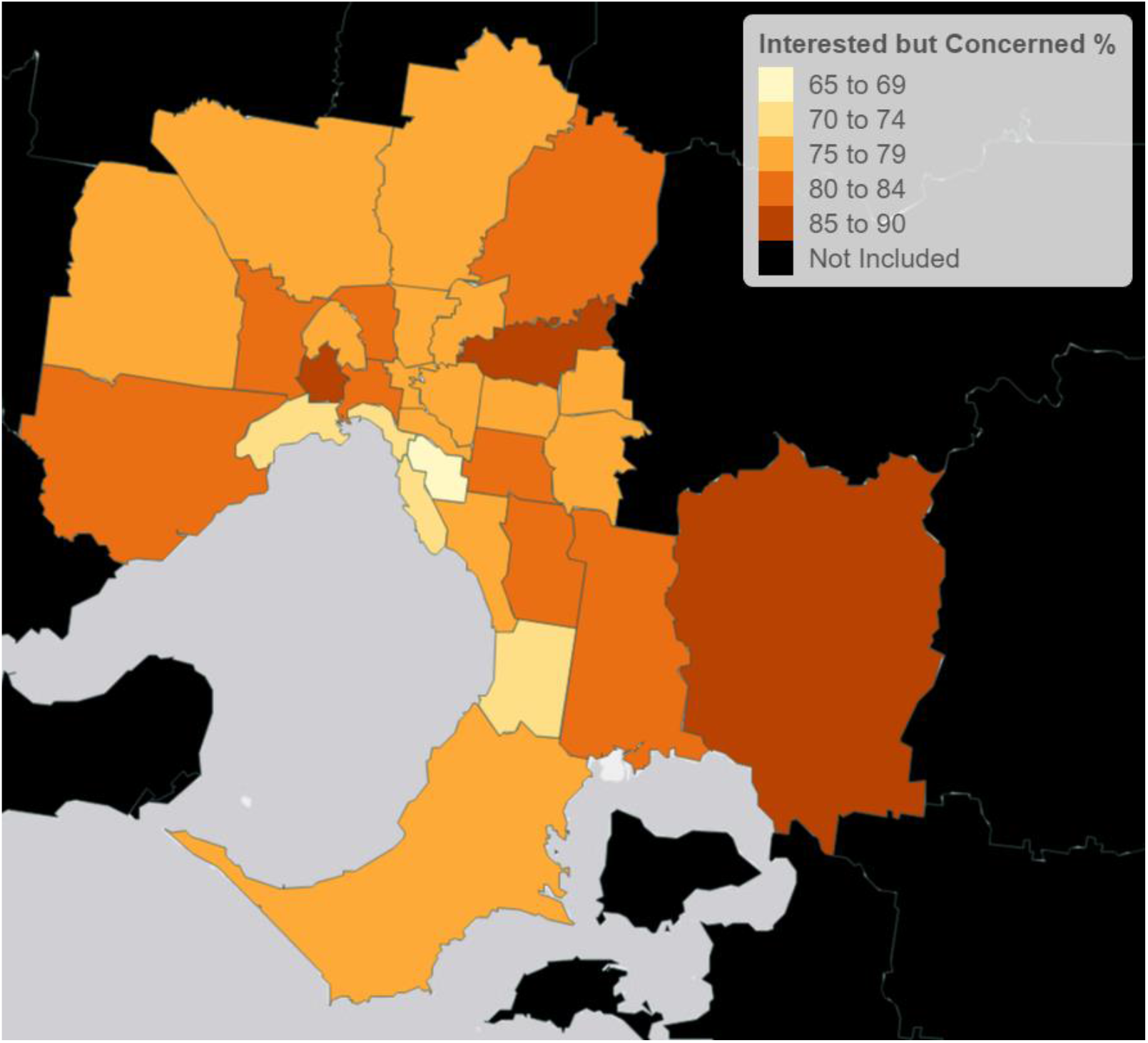
Heat map of Greater Melbourne showing the proportion of participants categorised as Interested but Concerned

Bike ownership was highest in the inner-West (67%), and lowest in inner-East areas, where in one LGA, only 40% of participants owned a bike. Participants who rode a bike at least once per week were most common in the City of Melbourne (32%), and generally decreased with distance from the city (Figure 6).

**Figure 6.**
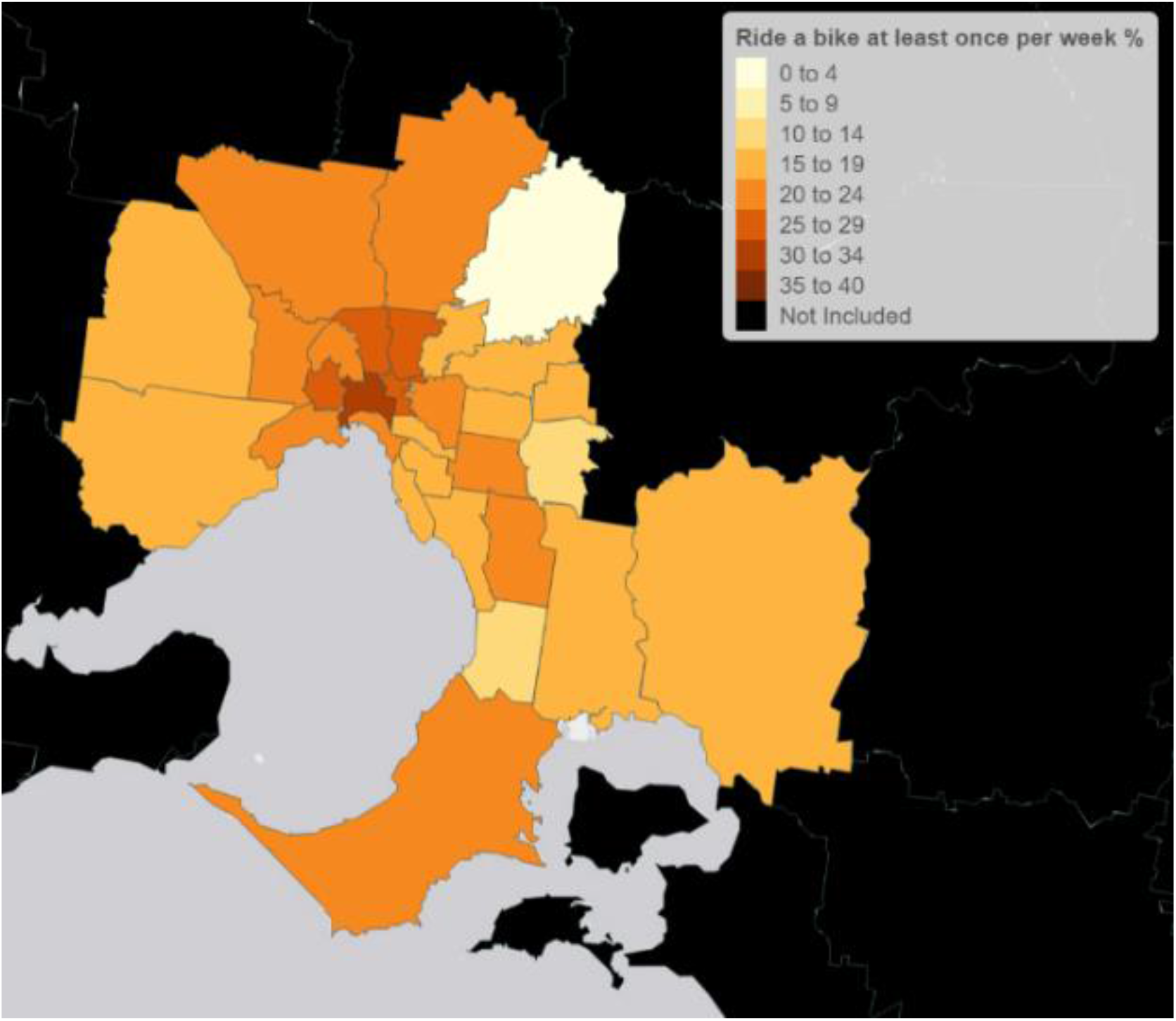
Heat map of Greater Melbourne showing the proportion of participants who rode a bike at least once per week prior to COVID-19 restrictions

### Regional comparisons

Proportions of Geller groups differed significantly between Greater Melbourne and regional Victorian LGAs (*x*^2^ = 19.4, *p* = <0.001). For both Greater Melbourne and regional Victorian LGAs, the Interested but Concerned group made up the majority of the sample (79% and 71% respectively). A higher proportion of participants were classified as No Way No How in regional Victoria (21%) than Greater Melbourne (15%), however bike ownership was comparable between regional Victoria (60%) and Greater Melbourne (57%). There was no association between how often participants rode a bike based on regionality (*x*^2^ = 0.5, *p* = 3.5) (Supplementary Material C).

### Age Comparisons

More than half of each sample were Interested but Concerned in all age groups (Figure 7). There was an association between age group and the Geller distribution (*x*^2^ = 214.7, *p* = <0.001). The proportion of participants classified as Interested but Concerned was lower in participants aged 55-74 years (72%) and 75 or more years (55%) compared to participants aged 18-34 years (84%) and 35 to 54 years (83%). The proportion of participants classified as No Way No How was highest in participants aged 75 years and older (42%) and 55-74 years (23%).

**Figure 7.**
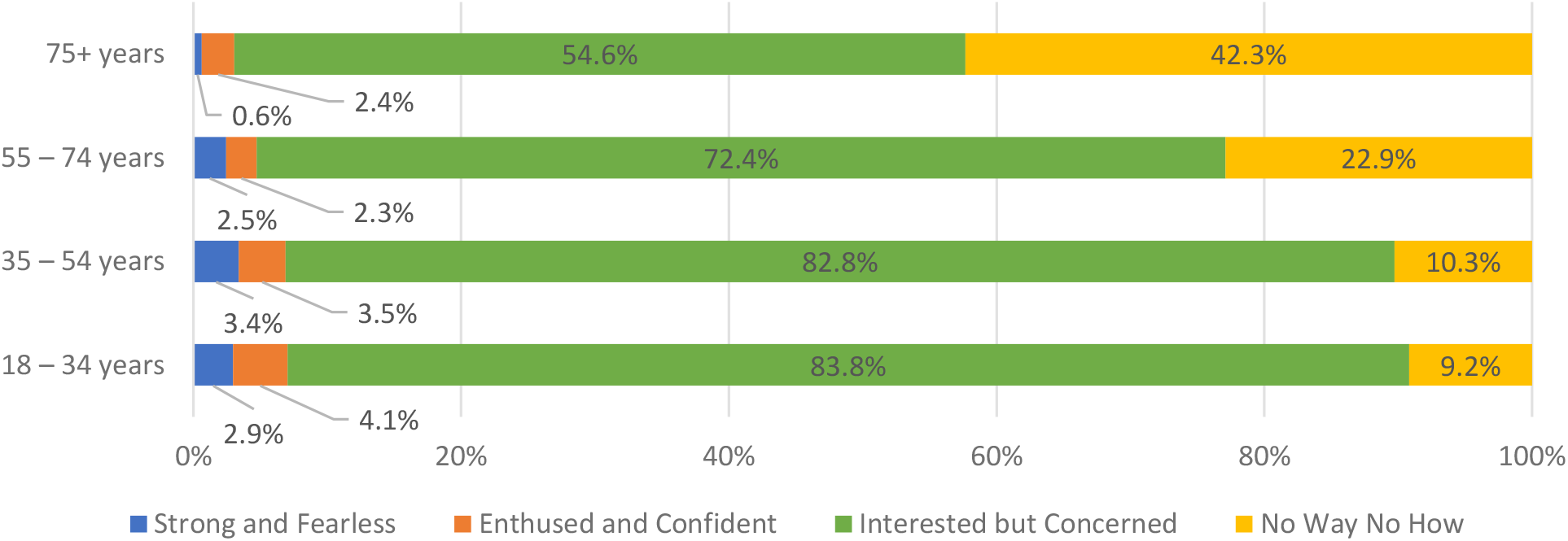
Distribution of Geller groups within age categories (weighted)

### Gender comparisons

A higher proportion of men than women in the sample owned a bike (63% vs. 52%), rode a bike at least once per week (28% vs. 12%) and rode a bike solely for transport (8.3% vs. 5.9%). The distribution of Geller groups differed between women and men (*x*^2^=79.7, *p*=<0.001) (see Figure 8). While the proportion of No Way No How participants was higher in women (19% vs 12%) and the proportion of Strong and Fearless (1.6% vs 4.0%) and Enthused and Confident (1.6% vs 4.7%) participants were lower in women, the proportion classified as Interested but Concerned was comparable in both women (78%) and men (79%).

**Figure 8.**
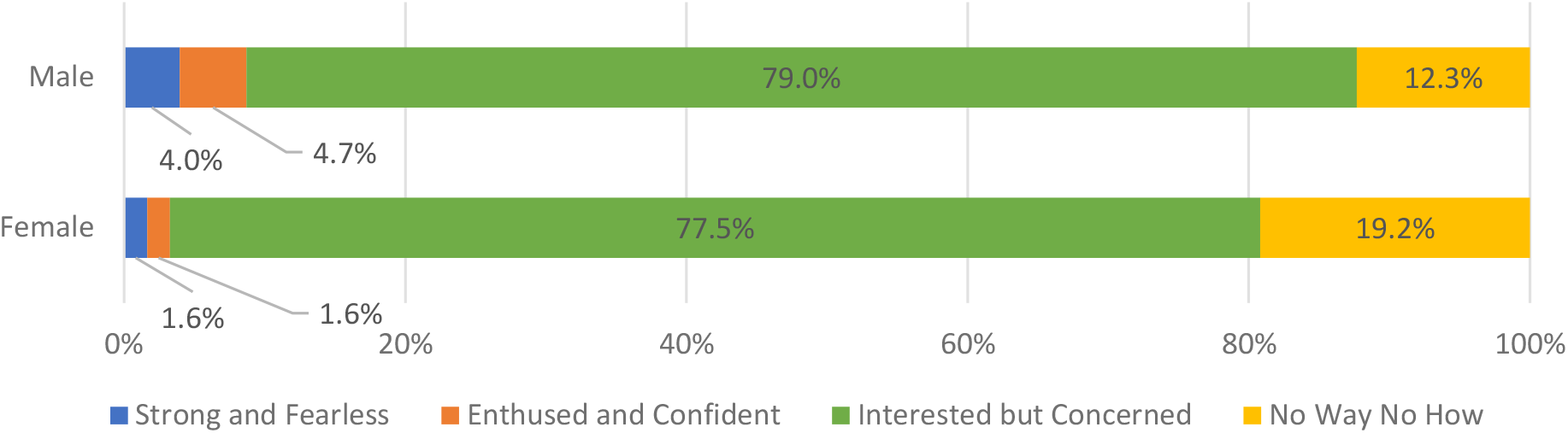
Distribution of Geller groups by gross income group (weighted)

### Income comparisons

A higher proportion of participants in the lower income groups ($1 - $20,799/year and $20,800 - $41,599/year) rode a bike more for transport, and a greater proportion rode four or more days per week than any other income groups. High income groups had the highest proportion of recreational bike riding (78% in participants earning $104,000 or more per year), and highest proportion of participants riding a bike 1-3 times per week (7.1% in $65,000 - $103,999/year group).

The proportion of Interested but Concerned participants ranged from 72% in the $20,800 - $41,599 income group to 82% in the $41,600 - $64,999 income group. The highest proportion of Strong and Fearless participants (4.2%) was in the group earning $104,000 per year or more (Figure 8 and Supplementary Material C).

**Figure 8a.**
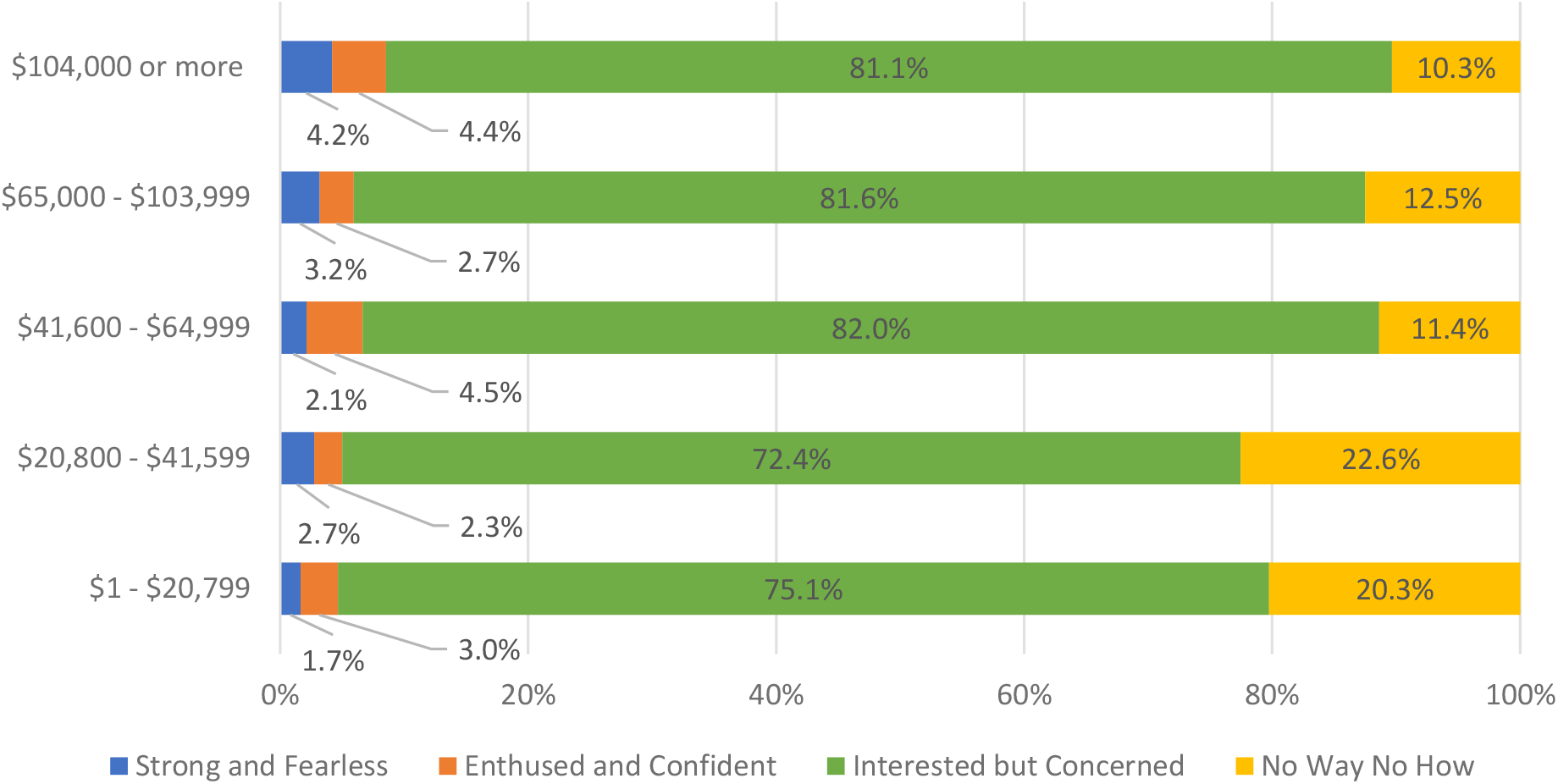
Distribution of Geller groups by gender (weighted)

## DISCUSSION

There is currently no data available on how interest in riding a bike differs between geographical regions, and no data on the Geller typologies in Australia. This study quantified the proportion of each Geller typology group across 37 LGAs in Victoria, Australia with complete data from 3999 individuals. Over half of participants owned a bike, however only one in five rode a bike at least once per week. Most participants were classified as Interested but Concerned, demonstrating a high latent demand for bike riding if protected bicycling infrastructure were provided. Geller group distributions differed between LGAs within Greater Melbourne, and between regional and metropolitan areas, supporting the need for analysis of Geller groups at smaller spatial scales than cities or states.

Few studies have used original research to identify the Geller typology proportions (8-11). Two sentinel Geller papers from the US (7, 8), one of which is published research, are often extrapolated to local government and state planning strategies in other areas (6, 31-34). Original research to identify Geller typology proportions has been conducted in the United States; including Portland (8, 9), Austin (9), Chicago (9), San Francisco (9), Washington D.C. (9), and in Edmonton, Canada (10). At times, these findings are applied to settings outside of where the data were collected. For example, the original paper from Roger Geller (7), based in Portland in the United States, has been cited in the Victorian Cycling Strategy in Australia (6). Research conducted in this area suggests 58% of people living in Portland are Interested but Concerned (8). Our Victorian-specific data demonstrates a far higher proportion classified as Interested but Concerned (78%).

Previous published studies using the Geller Typology to quantify interest in bike riding have done so at a multi-city, state and city level (8-11). To the authors’ knowledge, our study is the first to quantify the Geller typology proportions at a smaller spatial scale. We found that interest and confidence in riding a bike ranged from 66% to 88% within the metropolitan area of Greater Melbourne. These differences may come from differing available infrastructure, the demographics of an area, trip distance to central hubs and physical setting (e.g. topography, primary land-use type) (35, 36).

Further, it is important to identify these variations to understand the interest across areas with substantially different availability of bicycle infrastructure. While variation in Geller typologies was observed across Greater Melbourne, one of the most important findings was that interest in bike riding was consistently high (ranging from 66% to 88% across LGAs in Greater Melbourne), revealing a high latent demand across a vast geographical area of an entire metropolitan region. Currently, however, bicycling infrastructure is concentrated in inner-Melbourne to support higher volumes of bike traffic (12, 13). Therefore, bicycling infrastructure is not equitably provided across differing regions of Greater Melbourne (12, 13), likely contributing to both transport inequities (38), and health inequities through reduced physical activity participation (39) and a potentially increased risk of injury (40-44). The provision of protected bicycling infrastructure has the potential to address these inequities and support a potentially latent population of bike riders living in the outer urban fringe regions of Greater Melbourne (45-51). This would further enable current recommendations in Greater Melbourne of the “20-minute-neighbourhood” model, that supports people to meet most of their daily needs within a 20-minute trip from home (by walking, riding a bike or using public transport) (52). To inform the provision of locally-specific infrastructure, additional research is required to better understand trip purposes in outer urban fringe regions, and if they are focussed on transport to inner-city areas, or are for accessing local hubs and public transportation.

Women are often under-represented in bike riding campaigns and popular culture due to lower participation, and the perspective of bike riding being a male-dominated activity (17, 53, 54). Despite low participation, over two thirds of women in this study were interested in riding a bike and over half owned a bike. Importantly, many existing bike riding environments are often designed for the needs and confidence levels of men without disabilities (55-57). Infrastructure that enables women to ride a bike includes off-road paths, and bike-infrastructure that is physically separated from motor-vehicle traffic (55, 58). In future urban planning and research, it is vital that a lack of participation is not seen as a lack of interest in bike riding. Given that the findings of this study demonstrated high bike riding interest in women, understanding the specific barriers to riding a bike for women is needed to increase participation.

In this study, a higher proportion of people in the lower income categories rode a bike for transport purposes, and rode four or more days per week, compared to people in the higher income groups. Similar findings have been shown both in Australia and internationally, where there was an association between increased household income, and a decreased proportion of people riding a bike for transport (12, 59, 60). People in lower income groups may ride a bike out of necessity for commuting, rather than as a recreational activity. Despite this, people living in areas of lower socioeconomic status have disproportionately lower access to bicycling infrastructure (14, 61, 62). In Melbourne, much of the protected bicycling infrastructure are off-road paths located in parks, along rivers as trails, often in inner-city, higher socioeconomic areas (12, 13), with similar patterns internationally (14, 61-63). To support lower income groups in bicycle-commuting and reduce health inequities faced by low socioeconomic groups, high-quality and protected bicycling infrastructure should be provided equitably to support local travel and connections with public transport.

The provision of separated bicycling infrastructure is important for both the safety (64) and support of low risk bicycling environments to maintain and encourage participation. Most people in this study were interested in riding a bike if infrastructure were provided that physically separated them from motor vehicle traffic. While painted bike lanes are a lower cost alternative to providing bicycling infrastructure, these do not constitute physically separated bicycling infrastructure. Research conducted in Melbourne that measured passing distances between motor vehicles and bikes identified more close, and potentially unsafe, passes when a person riding a bike was travelling in a painted bike lane compared to on-road (65). Similarly, a previous study identified that 22% of all on-road bike riding crashes occurred while riding in a painted bike lane, highlighting their insufficiency in protecting vulnerable road-users (66). In addition to the risk of substantial injury that painted bike lanes pose for people on bikes, they are not supportive of new bike riders, or low-stress traffic environments, with concerns about safety on the road and interactions with motorists being a major barrier to participation in bike riding (17, 67-70). As indicated by the findings of this study, removing interactions with motor vehicles through a physically separated bicycle lane could substantially increase participation in bike riding in Melbourne, while maintaining the safety of vulnerable road-users.

To our knowledge, this was the first study of Geller typologies in Australia, and the largest and most population-representative for trip purpose and bike riding frequency. Further, this was the first application of the Geller typology at a small spatial scale, demonstrating important differences between local government areas. Despite this, some limitations were present. A recently published mixed methods study raised concerns over the internal validity of the Geller typology, particularly that the category participants were assigned to did not always represent their confidence in riding a bike (71). However, this typology was used because of its ease of understanding with policy makers, and to enable comparisons to existing literature and government bike riding strategies. Additionally, using an online research company may have favoured participants with a higher income, with computer access, and whom are able to regularly check their email, potentially introducing selection and response bias. To reduce response bias, invitations to complete the survey were sent in batches and at different times during the week. This enabled people who may have been occupied on certain days to receive a notification about the survey. As recommended by Hay et al., convenience internet panels are useful when data can be weighted to compensate for coverage errors (72). Some regional LGAs in this study had a low number of research company panel members. In turn, only a small number of participants completed surveys and represented the population in those LGAs. To reduce potential coverage errors, whole sample results were weighted by the population size of each LGA in an effort to balance the effects of under and over sampling across LGAs.

## CONCLUSION

To our knowledge, this study was the first application of the Geller typology groups at a small geographical area level. We demonstrated that while there was variation in interest in bike riding across an entire metropolitan region, interest was high across all LGAs, including outer urban areas that currently have lower participation in bike riding. Similarly, despite participation in bike riding being lower in women, interest remained high. It is vital that a lack of participation is not seen as a lack of interest in bike riding. These findings are important in considering the role that bike riding can play in reducing health and transport inequities and how protected bicycling infrastructure can be implemented to address these inequities. Further research is required to identify what factors are preventing people in Victoria from riding a bike and how these barriers vary across geographical areas.

## Data Availability

All data produced are available online at https://doi.org/10.1016/j.jth.2021.101290

## ACKNOWLEDGEMENTS

The authors would like to acknowledge the contribution of the participants in this study.

## REFERENCES

1. Oja P, Titze S, Bauman A, De Geus B, Krenn P, Reger-Nash B, et al. Health benefits of cycling: a systematic review. Scand J Med Sci Sports. 2011;21(4):496–509.

2. Warburton DE, Nicol CW, Bredin SS. Health benefits of physical activity: the evidence. Cmaj. 2006;174(6):801–9.

3. Rabl A, De Nazelle A. Benefits of shift from car to active transport. Transport policy. 2012;19(1):121–31.

4. Pérez K, Olabarria M, Rojas-Rueda D, Santamariña-Rubio E, Borrell C, Nieuwenhuijsen M. The health and economic benefits of active transport policies in Barcelona. Journal of Transport & Health. 2017;4:316–24.

5. Litman T. Evaluating active transport benefits and costs: Victoria Transport Policy Institute; 2015.

6. The Victorian Health Promotion Foundation. Victorian Cycling Strategy 2018-28. In: Department of Transport, Melbourne, Australia, 2017.

7. Geller R. Four Types of Cyclists. Portland, OR: Portland Office of Transportation; 2006.

8. Dill J, McNeil N. Four types of cyclists? Examination of typology for better understanding of bicycling behavior and potential. Transportation Research Record. 2013;2387(1):129–38.

9. McNeil N, Monsere CM, Dill J. Influence of bike lane buffer types on perceived comfort and safety of bicyclists and potential bicyclists. Transportation Research Record: National Research Council; 2015. p. 132–42.

10. Cabral L, Kim AM. An empirical reappraisal of the four types of cyclists. Transportation Research Part A: Policy and Practice. 2020;137:206–21.

11. Dill J, McNeil N. Revisiting the four types of cyclists: findings from a national survey. Transportation research record. 2016;2587(1):90–9.

12. Pistoll C, Goodman A. The link between socioeconomic position, access to cycling infrastructure and cycling participation rates: An ecological study in Melbourne, Australia. Journal of Transport & Health. 2014;1(4):251–9.

13. Kamphuis C, Giskes K, Kavanagh AM, Thornton L, Thomas LR, van Lenthe FJ, et al. Area variation in recreational cycling in Melbourne: a compositional or contextual effect? J Epidemiol Community Health. 2008;62(10):890–8.

14. Braun LM, Rodriguez DA, Gordon-Larsen P. Social (in) equity in access to cycling infrastructure: Cross-sectional associations between bike lanes and area-level sociodemographic characteristics in 22 large US cities. Journal of transport geography. 2019;80:102544.

15. Health and Human Services, Victorian State Government. Victoria’s restriction levels. 2020.

16. VicHealth. Values-based messaging for health promotion n.d. [Available from: https://www.vichealth.vic.gov.au/media-and-resources/hpcomms.

17. Daley M, Rissel C. Perspectives and images of cycling as a barrier or facilitator of cycling. Transport policy. 2011;18(1):211–6.

18. Australian Bureau of Statistics. National, state and territory population 2020 [Available from: https://www.abs.gov.au/statistics/people/population/national-state-and-territory-population/latest-release#states-and-territories.

19. Australian Bureau of Statistics. 2016 Census QuickStats - Greater Melbourne. 2016.

20. Victorian Inegrated Survey of Travel & Activity (VISTA). Total journeys to work by region and mode. 2018.

21. Australian Bureau of Statistics. 1270.0.55.003 - Australian Statistical Geography Standard (ASGS): Volume 3 - Non ABS Structures, July 2016.

22. Vic Councils. Roads & transport. n.d.

23. Qualtrics. Qualtrics. Provo, Utah, USA 2005.

24. Australian Bureau of Statistics. Australian Statistical Geographical Standard (ASGS). 2011.

25. Australian Bureau of Statistics. 2016 Census Canberra, ACT, Australia 2016 [Available from: https://www.abs.gov.au/websitedbs/censushome.nsf/home/2016.

26. RStudio Team. RStudio: Integrated Development for R.. Boston, MA: RStudio, PBC; 2020.

27. Lumley T. Package ’Survey.’ 2015 [Available from: http://cran.r-project.org/web/packages/survey/survey.pdf.

28. Australian Bureau of Statistics. Census of Population and Housing, 2016, 2011 and 2006. 2018.

29. Australian Bureau of Statistics. 3218.0 Regional Population Growth, Australia, 2017-2018.2019.

30. Graul C. leafletR: Interactive Web - Maps Based on the Leaflet JavaScript Library. R package version 0.4-0, 2016.

31. Transportation CDo. Chicago Streets for Cycling Plan 2020. Chicago Department of Transportation Chicago IL; 2012.

32. Olson J, Goff P, Piper S, Zeftling L, Buffalo G. Buffalo Bicycle Master Plan Update. New York State Energy Research and Development Authority; 2016.

33. City of Burlington. Cycling Master Plan. 2009.

34. Mississaug. Mississauga Cycling Master Plan. 2019.

35. Winters M, Brauer M, Setton EM, Teschke K. Built environment influences on healthy transportation choices: bicycling versus driving. J Urban Health. 2010;87(6):969–93.

36. Winters M, Brauer M, Setton EM, Teschke K. Mapping bikeability: a spatial tool to support sustainable travel. Environment and Planning B: Planning and Design. 2013;40(5):865–83.

37. Nykiforuk CI, Flaman LM. Geographic information systems (GIS) for health promotion and public health: a review. Health Promot Pract. 2011;12(1):63–73.

38. Lee RJ, Sener IN, Jones SN. Understanding the role of equity in active transportation planning in the United States. Transport reviews. 2017;37(2):211–26.

39. Goodman A, Sahlqvist S, Ogilvie D, iConnect C. New walking and cycling routes and increased physical activity: one- and 2-year findings from the UK iConnect Study. Am J Public Health. 2014;104(9):e38–46.

40. Cicchino JB, McCarthy ML, Newgard CD, Wall SP, DiMaggio CJ, Kulie PE, et al. Not all protected bike lanes are the same: Infrastructure and risk of cyclist collisions and falls leading to emergency department visits in three US cities. Accident Analysis & Prevention. 2020;141:105490.

41. Reynolds CC, Harris MA, Teschke K, Cripton PA, Winters M. The impact of transportation infrastructure on bicycling injuries and crashes: a review of the literature. Environmental health. 2009;8(1):1–19.

42. Teschke K, Frendo T, Shen H, Harris MA, Reynolds CC, Cripton PA, et al. Bicycling crash circumstances vary by route type: a cross-sectional analysis. BMC public health. 2014;14(1):1–10.

43. Teschke K, Harris MA, Reynolds CC, Winters M, Babul S, Chipman M, et al. Route infrastructure and the risk of injuries to bicyclists: a case-crossover study. Am J Public Health. 2012;102(12):2336–43.

44. Morrison CN, Thompson J, Kondo MC, Beck B. On-road bicycle lane types, roadway characteristics, and risks for bicycle crashes. Accident Analysis & Prevention. 2019;123:123–31.

45. Goodman A, Sahlqvist S, Ogilvie D, Consortium i. New walking and cycling routes and increased physical activity: one-and 2-year findings from the UK iConnect Study. Am J Public Health. 2014;104(9):e38–e46.

46. Panter J, Heinen E, Mackett R, Ogilvie D. Impact of new transport infrastructure on walking, cycling, and physical activity. American journal of preventive medicine. 2016;50(2):e45–e53.

47. Heinen E, Panter J, Mackett R, Ogilvie D. Changes in mode of travel to work: a natural experimental study of new transport infrastructure. International Journal of Behavioral Nutrition and Physical Activity. 2015;12(1):1–10.

48. Mölenberg FJ, Panter J, Burdorf A, van Lenthe FJ. A systematic review of the effect of infrastructural interventions to promote cycling: strengthening causal inference from observational data. International journal of behavioral nutrition and physical activity. 2019;16(1):1–31.

49. Prins RG, Panter J, Heinen E, Griffin SJ, Ogilvie DB. Causal pathways linking environmental change with health behaviour change: Natural experimental study of new transport infrastructure and cycling to work. Prev Med. 2016;87:175–82.

50. Smith M, Hosking J, Woodward A, Witten K, MacMillan A, Field A, et al. Systematic literature review of built environment effects on physical activity and active transport–an update and new findings on health equity. International journal of behavioral nutrition and physical activity. 2017;14(1):1–27.

51. Pucher J, Dill J, Handy S. Infrastructure, programs, and policies to increase bicycling: an international review. Prev Med. 2010;50:S106–S25.

52. Department of Environment L, Water and Planning. Plan Melbourne 2017-2050. In: Department of Environment L, Water and Planning, Victorian State Government, editor. Melbourne, Australia 2017.

53. Bauman AE, Blazek K, Reece L, Bellew W. The emergence and characteristics of the Australian Mamil. Med J Aust. 2018;209(11):490–4.

54. Knight A. Cycling is the new golf. Sydney Morning Herald. 2014.

55. Aldred R, Elliott B, Woodcock J, Goodman A. Cycling provision separated from motor traffic: a systematic review exploring whether stated preferences vary by gender and age. Transport reviews. 2017;37(1):29–55.

56. Mitra R, Nash S. Does the Built Environment Explain Gender Gap in Cycling? a Study of Post-Secondary Students in Toronto, Canada. Journal of Transport & Health. 2017;5:S24–S5.

57. Xie L, Spinney J. “I won’t cycle on a route like this; I don’t think I fully understood what isolation meant": A critical evaluation of the safety principles in Cycling Level of Service (CLoS) tools from a gender perspective. Travel behaviour and society. 2018;13:197–213.

58. Garrard J, Rose G, Lo SK. Promoting transportation cycling for women: the role of bicycle infrastructure. Prev Med. 2008;46(1):55–9.

59. Heesch KC, Giles-Corti B, Turrell G. Cycling for transport and recreation: associations with socio-economic position, environmental perceptions, and psychological disposition. Prev Med. 2014;63:29–35.

60. Xing Y, Handy SL, Mokhtarian PL. Factors associated with proportions and miles of bicycling for transportation and recreation in six small US cities. Transportation research part D: Transport and Environment. 2010;15(2):73–81.

61. Mora R, Truffello R, Oyarzún G. Equity and accessibility of cycling infrastructure: An analysis of Santiago de Chile. Journal of Transport Geography. 2021;91:102964.

62. Flanagan E, Lachapelle U, El-Geneidy A. Riding tandem: Does cycling infrastructure investment mirror gentrification and privilege in Portland, OR and Chicago, IL? Research in Transportation Economics. 2016;60:14–24.

63. Tucker B, Manaugh K. Bicycle equity in Brazil: Access to safe cycling routes across neighborhoods in Rio de Janeiro and Curitiba. International journal of sustainable transportation. 2018;12(1):29–38.

64. Harris MA, Reynolds CC, Winters M, Cripton PA, Shen H, Chipman ML, et al. Comparing the effects of infrastructure on bicycling injury at intersections and non-intersections using a case– crossover design. Injury prevention. 2013;19(5):303–10.

65. Beck B, Chong D, Olivier J, Perkins M, Tsay A, Rushford A, et al. How much space do drivers provide when passing cyclists? Understanding the impact of motor vehicle and infrastructure characteristics on passing distance. Accident Analysis & Prevention. 2019.

66. Beck B, Stevenson M, Newstead S, Cameron P, Judson R, Edwards ER, et al. Bicycling crash characteristics: An in-depth crash investigation study. Accident Analysis & Prevention. 2016;96:219–27.

67. Heesch KC, Sahlqvist S, Garrard J. Gender differences in recreational and transport cycling: a cross-sectional mixed-methods comparison of cycling patterns, motivators, and constraints. International Journal of Behavioral Nutrition and Physical Activity. 2012;9(1):106.

68. Twaddle H, Hall F, Bracic B. Latent bicycle commuting demand and effects of gender on commuter cycling and accident rates. Transportation Research Record2010. p. 28–36.

69. Akar G, Clifton KJ. Influence of individual perceptions and bicycle infrastructure on decision to bike. Transportation Research Record2009. p. 165–72.

70. Dill J. Bicycling for transportation and health: the role of infrastructure. J Public Health Policy. 2009;30 Suppl 1:S95–110.

71. Hosford K, Laberee K, Fuller D, Kestens Y, Winters M. Are they really interested but concerned? A mixed methods exploration of the Geller bicyclist typology. Transportation Research Part F: Traffic Psychology and Behaviour. 2020;75:26–36.

72. Hays RD, Liu H, Kapteyn A. Use of Internet panels to conduct surveys. Behavior research methods. 2015;47(3):685–90.

