## Supplementary Materials for "The potential for bike riding across entire cities: quantifying spatial variation in interest in bike riding"

**Supplementary Material A - Study Survey**

Q1. What is your residential postcode? (if postcode that is on LGA boundary – referred to map)

Q2. What is your age?

- 18-24
- 25-34
- 35-44
- 45-64
- 65-74
- 75+

Q3. What gender do you identify as?

- Female
- Male
- Non-binary
- Prefer not to say
- Other (please specify)

Q4. Please indicate the answer that includes your personal income in 2019 before taxes.

- $1 to $10,399
- $10,400 to $20,799
- $20,800 to $31,199
- $31,200 to $41,599
- $41,600 to $51,999
- $52,000 to $64,999
- $65,000 to $77,999
- $78,000 to $103,999
- $104,000 or more

Q5. Is English the main language you speak at home?

- Yes
- No (please specify)

Q6. Do you own a bike?

- Yes
- No

Q7. Do you have an injury or condition that prevents you from riding a bike?

- Yes
- No

Q8. Prior to COVID-19 restrictions, had you ridden a bike in the last 12 months?

- Yes
- No

*If "Prior to COVID-19 restrictions, had you ridden a bike in the last 12 months?" = Yes, directed to:*

Q8.1. Prior to COVID-19 restrictions, had you ridden a bike in the last month?

- Yes
- No

*If "Prior to COVID-19 restrictions, had you ridden a bike in the last month?" = Yes, directed to:*

Q8.2. Have you continued to ride a bike during COVID-19?

- Yes
- No

*If "Prior to COVID-19 restrictions, had you ridden a bike in the last 12 months?" = No OR ""Prior to COVID-19 restrictions, had you ridden a bike in the last month?" = No, directed to:*

Q8.3. Have you begun riding a bike (or begun riding again) COVID-19?

- Yes

- No

Q9. Do you have any interest in riding a bike when COVID-19 restrictions are lifted?

- Yes
- Maybe
- No

Q10. Prior to COVID-19, how often did you ride a bike?

- Never
- 1-3 times per year
- 1-3 times per month
- 1-3 times per week
- 4+ days per week

*If "Prior to COVID-19, how often did you ride a bike?" was not "Never", directed to:*

Q10.1. When you ride a bike, for what purpose do you do so?

- Transport (e.g. to get to the shops, to work)
- Recreation (e.g. for exercise)
- Both transport and recreation

This section will ask you about how comfortable you would feel riding a bike in different settings.

Q11. How comfortable would you feel riding a bike on a path or trail separate from the street?

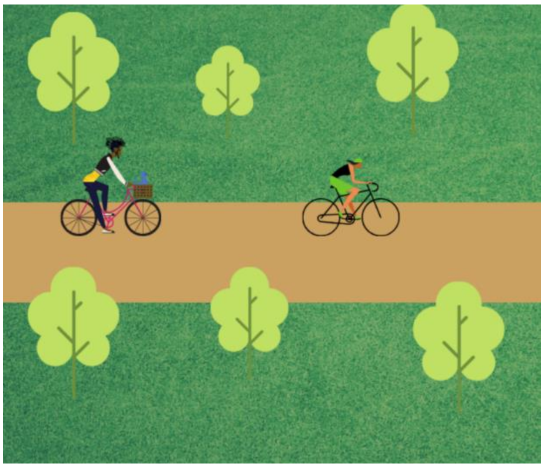

- Sliding scale – 1 (Very Uncomfortable) to 4 (Very Comfortable)

Q12. How comfortable would you feel riding a bike on a quiet, residential street with traffic speeds of 30-40 km per hour?

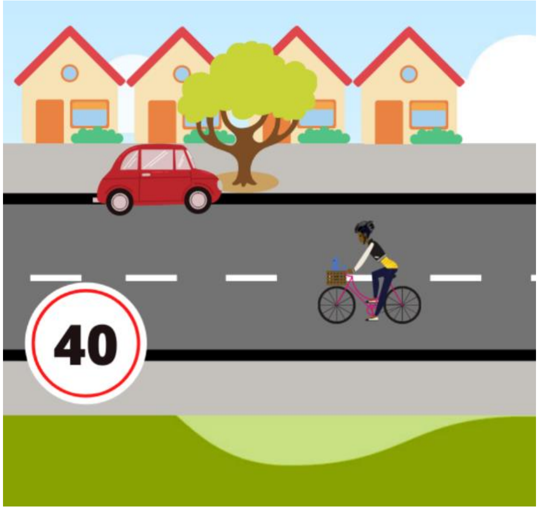


- Sliding scale – 1 (Very Uncomfortable) to 4 (Very Comfortable)

Q13. How comfortable would you feel on a quiet residential street with bicycle route markings, wide speed humps, and other things that slow down and discourage car traffic?

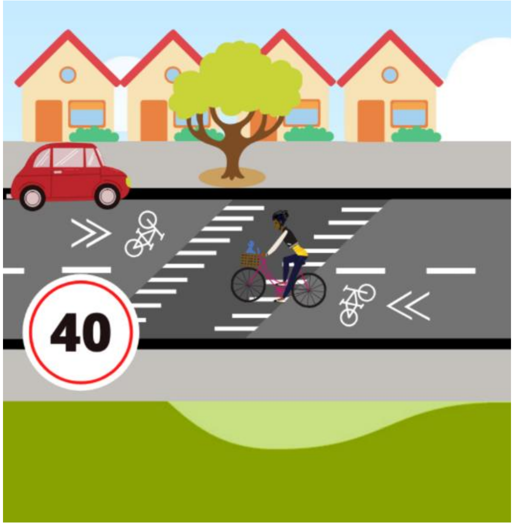


- Sliding scale – 1 (Very Uncomfortable) to 4 (Very Comfortable)

Q14. How comfortable would you feel riding a bike on a two-lane commercial shopping street with traffic speeds of 30-40 km/h, on-street parking and no bike lane?

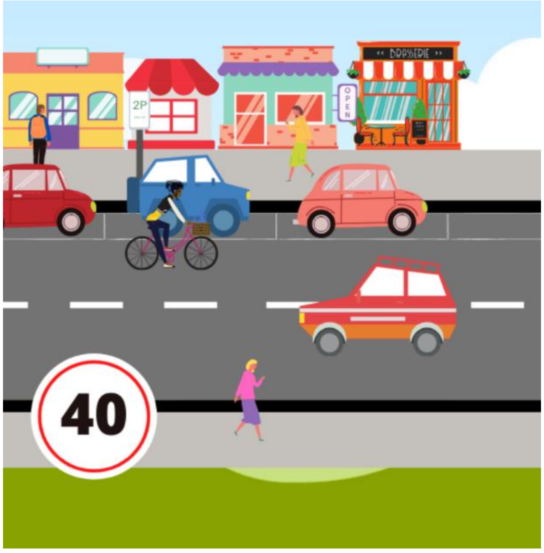


- Sliding scale – 1 (Very Uncomfortable) to 4 (Very Comfortable)

Q15. How comfortable would you feel riding a bike on a two-lane commercial shopping street with traffic speeds of 30-40 km/h, on-street parking and a painted bike-lane?

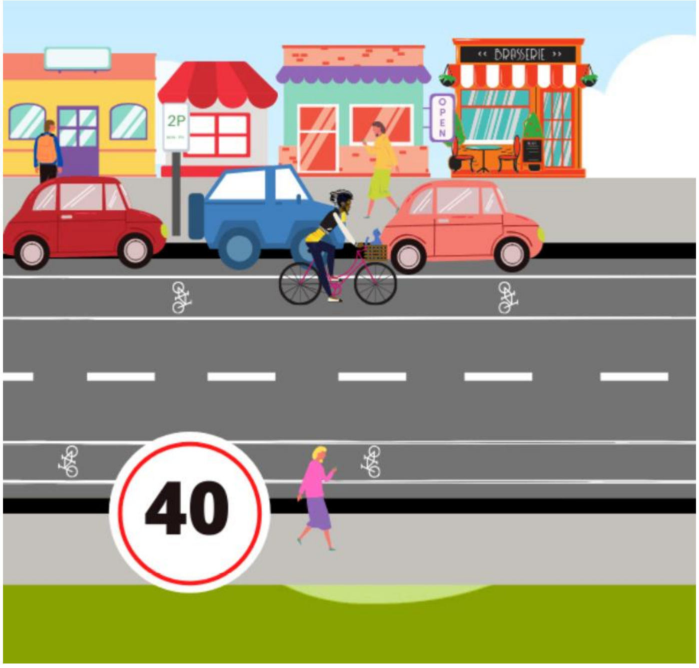


- Sliding scale – 1 (Very Uncomfortable) to 4 (Very Comfortable)

Q16. How comfortable would you feel riding a bike on a major urban or suburban street with four lanes, on-street parking, traffic speeds of 50-60 km/h, and no bike lane?

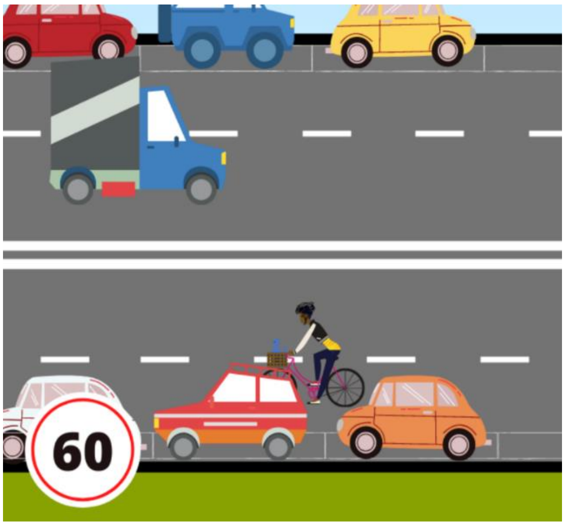


- Sliding scale – 1 (Very Uncomfortable) to 4 (Very Comfortable)

Q17. How comfortable would you feel riding a bike on a major urban or suburban street with four lanes, on-street parking, traffic speeds of 50-60 km/h, and a painted bike-lane?

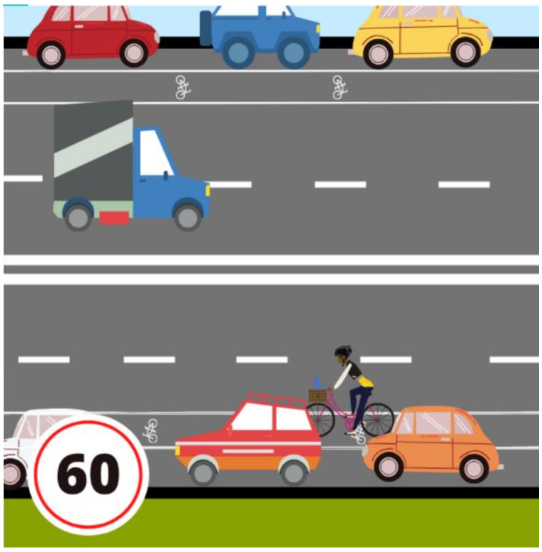


- Sliding scale – 1 (Very Uncomfortable) to 4 (Very Comfortable)

Q18. How comfortable would you feel riding a bike on a major urban or suburban street with four lanes, on-street parking, traffic speeds of 50-60 km/h, and a wide bike lane separated from traffic by a raised curb or parked cars?

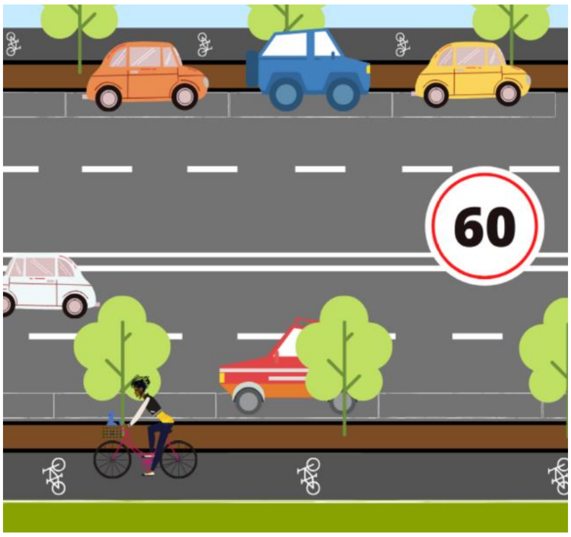


- Sliding scale – 1 (Very Uncomfortable) to 4 (Very Comfortable)

Q19. How comfortable would you feel riding a bike on a major street with two lanes in each direction, a centre divider, on-street parking, traffic speeds 50-60 km/h, and no bike lane?

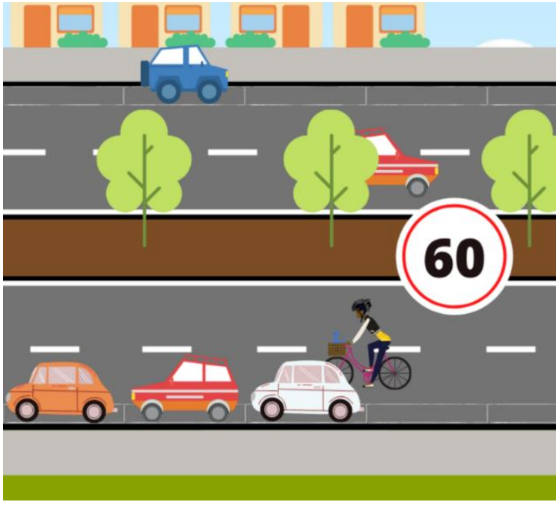


- Sliding scale – 1 (Very Uncomfortable) to 4 (Very Comfortable)

Q20. How comfortable would you feel riding a bike on a major street with two lanes in each direction, a centre divider, on-street parking, traffic speeds 50-60 km/h, and a wide bike lane separated from traffic by a raised curb or parked cars?

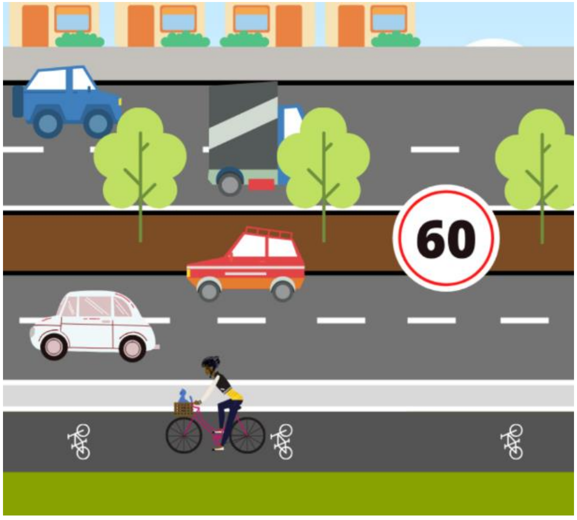


- Sliding scale – 1 (Very Uncomfortable) to 4 (Very Comfortable)

Q21. Please indicate how you feel about the following statement: "I would like to ride a bike more"

- Sliding scale – 1 (Strongly Disagree) to 4 (Strongly Agree)

### Supplementary Material B – Demographic characteristics

*Table 1. Calculated sample sizes for each LGA based on description in methods, the total number of ORU panel members residing in each LGA (member panel total) and the number of people who then completed a survey and were included in the study (total sample size)*

| Local Government Area | Recommended Sample Size (*n*) | Member Panel Total (*n*) | Total Sample Size (*n*) |
|---|---|---|---|
| ***Greater Melbourne LGAs*** | | | |
| Banyule City Council | 79 | 165 | 85 |
| Bayside City Council | 64 | 106 | 88 |
| Brimbank City Council | 127 | 294 | 136 |
| Cardinia Shire Council | 65 | 103 | 75 |
| City of Boroondara | 110 | 194 | 123 |
| City of Casey | 207 | 276 | 229 |
| City of Darebin | 98 | 185 | 102 |
| City of Glen Eira | 94 | 195 | 98 |
| City of Greater Dandenong | 101 | 145 | 122 |
| City of Kingston | 99 | 147 | 121 |
| City of Knox | 99 | 133 | 129 |
| City of Melbourne | 103 | 450 | 152 |
| City of Melton | 95 | 205 | 131 |
| City of Monash | 122 | 205 | 143 |
| City of Moonee Valley | 78 | 144 | 112 |
| City of Port Phillip | 69 | 251 | 73 |
| City of Stonnington | 71 | 166 | 86 |
| City of Whitehorse | 107 | 173 | 158 |
| City of Whittlesea | 136 | 221 | 145 |
| City of Yarra | 60 | 112 | 74 |
| Frankston City Council | 86 | 151 | 103 |
| Hobsons Bay City Council | 59 | 95 | 67 |
| Hume City Council | 136 | 168 | 168 |
| Manningham Council | 76 | 128 | 126 |
| Maribyrnong City Council | 56 | 120 | 72 |
| Maroondah City Council | 71 | 119 | 99 |

| | | | |
|---|---|---|---|
| Moreland City Council | 110 | 236 | 147 |
| Mornington Peninsula Shire | 101 | 98 | 110 |
| Nillumbik Shire Council | 39 | 101 | 41 |
| Wyndham City | 155 | 203 | 197 |
| Yarra Ranges Shire | 96 | 138 | 102 |
| ***Regional LGAs*** | | | |
| City of Ballarat | 63 | 104 | 75 |
| City of Greater Bendigo | 68 | 83 | 80 |
| City of Greater Geelong | 149 | 185 | 166 |
| City of Latrobe | 44 | 53 | 60 |
| Greater Shepparton City Council | 39 | 44 | 48 |
| Surf Coast Shire | 19 | 35 | 21 |

**Supplementary Material C – Demographic characteristics**

*Table 3. Demographic characteristics of the weighted sample and target population*

| | Unweighted Study Sample (*n* = 3999) | Weighted Study Sample (*n* = 3999) | Aggregated LGA Population* |
|---|---|---|---|
| **Gender** | | | |
| Female | 48.8% | 49.1% | 51.0% |
| Male | 50.7% | 50.5% | 49.0% |
| Non-binary | 0.3% | 0.3% | N/A |
| Other | 0.0% | 0.0% | N/A |
| Prefer not to say | 0.2% | 0.1% | N/A |
| **Age Category** | | | |
| 18-24 years | 11.2% | 11.1% | 12.7% |
| 25-34 years | 18.9% | 18.7% | 20.3% |
| 35-44 years | 16.0% | 15.7% | 17.9% |
| 45-54 years | 15.5% | 15.5% | 16.7% |
| 55-64 years | 17.7% | 17.9% | 13.9% |
| 65-74 years | 16.7% | 16.9% | 10.1% |
| 75+ years | 4.1% | 4.2% | 8.4% |
| **Income** | | | |
| $1 - $10,399 | 8.3% | 8.3% | 9.4% |
| $10,400 - $20,799 | 8.7% | 8.6% | 23.2% |
| $20,800 - $31,199 | 11.5% | 11.6% | 13.3% |
| $31,200 - $41,599 | 8.9% | 9.0% | 12.4% |
| $41,600 - $51,999 | 10.6% | 10.6% | 10.3% |
| $52,000 - $64,999 | 11.3% | 11.3% | 9.8% |
| $65,000 - $77,999 | 10.4% | 10.3% | 6.8% |
| $78,000 - $103,999 | 15.5% | 15.5% | 7.5% |
| $104,000 + | 14.8% | 14.6% | 7.4% |
| **English not main language at home** | 12.0% | 11.8% | 22.6%** |
| **English main language at home** | | 88.2% | 77.4% |

*based on ABS Census 2016 data (25)

**in all of Victoria (28)

**Supplementary Material D – Proportions and chi-square results for region of residence, gender, age and income**

*Table 4. Bike-riding behaviours and Geller Groups based on region of residence*

| | Greater Melbourne LGAs | Regional LGAs | *p*-value | $X^2$ | *f* |
|---|---|---|---|---|---|
| **Proportion of weighted sample** | 88.1% | 11.9% | | | |
| **Geller groups** | | | <0.001 | 19.4 | 3 |
| Strong and Fearless | 2.7% | 3.4% | | | |
| Enthused and Confident | 3.0% | 4.6% | | | |
| Interested but Concerned | 79.3% | 70.5% | | | |
| No Way No How | 14.9% | 21.4% | | | |
| **Bike ownership** | | | 0.25 | 1.4 | 1 |
| Yes | 57.0% | 59.8% | | | |
| No | 43.0% | 40.2% | | | |
| **Frequency of riding a bike** | | | 0.51 | 3.5 | 4 |
| Never | 32.5% | 35.6% | | | |
| 1 - 3 times per year | 26.9% | 27.9% | | | |
| 1 - 3 times per month | 20.3% | 19.2% | | | |
| 1 - 3 times per week | 14.7% | 12.4% | | | |
| 4 + days per week | 5.6% | 4.8% | | | |
| **Primary trip purpose *** | | | 0.15 | 5.7 | 3 |
| Transport only | 7.4% | 6.3% | | | |
| Recreation only | 71.1% | 76.5% | | | |
| Both transport and recreation | 21.6% | 17.1% | | | |

* based on those who had ridden a bike in the 12 months previous to COVID-19 restrictions

*Table 5. Bike-riding behaviours and Geller Groups based on gender*

| | **Women** | **Men** | ***p*-value** | $X^2$ | ***f*** |
|---|---|---|---|---|---|
| **Proportion of weighted sample** | 49.3% | 50.7% | | | |
| **Geller groups** | | | <0.001 | 79.7 | |
| Strong and Fearless | 1.6% | 4.0% | | | 3 |
| Enthused and Confident | 1.6% | 4.7% | | | |
| Interested but Concerned | 77.5% | 79.0% | | | |
| No Way No How | 19.2% | 12.3% | | | |
| **Bike ownership** | | | <0.001 | 49.6 | 1 |
| Yes | 51.7% | 62.7% | | | |
| No | 48.3% | 37.3% | | | |
| **Frequency of riding a bike** | | | <0.001 | 199.8 | 4 |
| Never | 40.3% | 25.7% | | | |
| 1 - 3 times per year | 29.1% | 25.1% | | | |
| 1 - 3 times per month | 18.6% | 21.7% | | | |
| 1 - 3 times per week | 9.4% | 19.4% | | | |
| 4 + days per week | 2.7% | 8.2% | | | |
| **Primary trip purpose *** | | | <0.001 | 134.9 | 3 |
| Transport only | 5.9% | 8.3% | | | |
| Recreation only | 77.9% | 66.9% | | | |
| Both transport and recreation | 16.2% | 24.8% | | | |

* based on those who had ridden a bike in the 12 months previous to COVID-19 restrictions

*Table 6. Bike-riding behaviours and Geller Groups based on age group*

| | 18 – 34 years | 35 – 54 years | 55 – 74 years | 75+ years | *p*-value | $X^2$ | *f* |
|---|---|---|---|---|---|---|---|
| **Proportion of weighted sample** | 29.6% | 31.2% | 34.9% | 4.2% | | | |
| **Geller groups** | | | | | <0.001 | 214.7 | 9 |
| Strong and Fearless | 2.9% | 3.4% | 2.5% | 0.6% | | | |
| Enthused and Confident | 4.1% | 3.5% | 2.3% | 2.4% | | | |
| Interested but Concerned | 83.8% | 82.8% | 72.4% | 54.6% | | | |
| No Way No How | 9.2% | 10.3% | 22.9% | 42.3% | | | |
| **Bike ownership** | | | | | <0.001 | 167.4 | 3 |
| Yes | 64.0% | 64.0% | 50.0% | 20.4% | | | |
| No | 36.0% | 36.0% | 50.0% | 79.6% | | | |
| **Frequency of riding a bike** | | | | | <0.001 | 202.6 | 12 |
| Never | 22.6% | 30.4% | 39.9% | 64.9% | | | |
| 1 - 3 times per year | 27.2% | 26.9% | 28.0% | 20.1% | | | |
| 1 - 3 times per month | 24.4% | 21.3% | 17.3% | 5.6% | | | |
| 1 - 3 times per week | 18.6% | 15.0% | 11.2% | 7.5% | | | |
| 4 + days per week | 7.2% | 6.4% | 3.7% | 1.8% | | | |
| **Primary trip purpose *** | | | | | <0.001 | 375.0 | 9 |
| Transport only | 13.6% | 5.6% | 2.1% | 5.1% | | | |
| Recreation only | 56.8% | 73.4% | 85.4% | 84.3% | | | |
| Both transport and recreation | 29.5% | 21.0% | 12.5% | 10.6% | | | |

* based on those who had ridden a bike in the 12 months previous to COVID-19 restrictions

*Table 7. Bike-riding behaviours and Geller Groups based on income group*

| | $1 - $20,799 | $20,800 - $41,599 | $41,600 - $64,999 | $65,000 - $103,999 | $104,000 + | *p*-value | $X^2$ | *f* |
|---|---|---|---|---|---|---|---|---|
| **Proportion of weighted sample** | 16.8% | 20.6% | 22.0% | 25.9% | 14.7% | | | |
| **Geller groups** | | | | | | <0.001 | 79.1 | 15 |
| Strong and Fearless | 1.7% | 2.7% | 2.1% | 3.2% | 4.2% | | | |
| Enthused and Confident | 3.0% | 2.3% | 4.5% | 2.7% | 4.4% | | | |
| Interested but Concerned | 75.1% | 72.4% | 82.0% | 81.6% | 81.1% | | | |
| No Way No How | 20.3% | 22.6% | 11.4% | 12.5% | 10.3% | | | |
| **Bike ownership** | | | | | | <0.001 | 99.7 | 5 |
| Yes | 51.5% | 47.0% | 61.1% | 65.3% | 67.4% | | | |
| No | 48.5% | 53.0% | 38.9% | 34.7% | 32.6% | | | |
| **Frequency of riding a bike** | | | | | | <0.001 | 133.9 | 20 |
| Never | 18.9% | 15.3% | 22.1% | 24.0% | 24.6% | | | |
| 1 - 3 times per year | 12.6% | 13.1% | 17.1% | 18.1% | 14.2% | | | |
| 1 - 3 times per month | 26.7% | 25.1% | 27.6% | 25.9% | 28.0% | | | |
| 1 - 3 times per week | 4.4% | 5.3% | 5.5% | 7.3% | 6.2% | | | |
| 4 + days per week | 37.5% | 41.2% | 27.7% | 24.6% | 27.0% | | | |
| **Primary trip purpose *** | | | | | | | | |
| Transport only | 13.5% | 11.1% | 7.7% | 3.7% | 3.9% | <0.001 | 167.8 | 15 |
| Recreation only | 60.9% | 68.5% | 70.4% | 72.6% | 77.5% | | | |
| Both transport and recreation | 25.6% | 20.4% | 21.9% | 23.7% | 18.6% | | | |

* based on those who had ridden a bike in the 12 months previous to COVID-19 restrictions

**Supplementary Material D – Four Types of Cyclist proportions by LGA**

| | Strong and Fearless (%) | Enthused and Confident (%) | Interested but Concerned (%) | No Way No How (%) |
|---|---|---|---|---|
| Banyule City Council | 7.1 | 3.5 | 78.8 | 10.6 |
| Bayside City Council | 6.0 | 3.6 | 74.7 | 15.7 |
| Brimbank City Council | 2.2 | 0.7 | 83.6 | 13.4 |
| Cardinia Shire Council | 0.0 | 0.0 | 85.3 | 14.7 |
| City of Boroondara | 2.5 | 3.3 | 77.5 | 16.7 |
| City of Casey | 3.1 | 0.9 | 82.1 | 13.9 |
| City of Darebin | 2.0 | 3.0 | 75.0 | 20.0 |
| City of Glen Eira | 6.2 | 5.2 | 66.0 | 22.7 |
| City of Greater Dandenong | 0.8 | 5.7 | 81.1 | 12.3 |
| City of Kingston | 2.5 | 3.4 | 77.3 | 16.8 |
| City of Knox | 4.0 | 2.4 | 78.4 | 15.2 |
| City of Melbourne | 3.9 | 3.3 | 84.2 | 8.6 |
| City of Melton | 3.1 | 2.3 | 78.3 | 16.3 |
| City of Monash | 0.0 | 1.4 | 83.5 | 15.1 |
| City of Moonee Valley | 6.4 | 3.6 | 77.3 | 12.7 |
| City of Port Phillip | 2.8 | 4.2 | 70.8 | 22.2 |
| City of Stonnington | 0.0 | 1.2 | 78.8 | 20.0 |
| City of Whitehorse | 0.6 | 4.5 | 77.6 | 17.3 |
| City of Whittlesea | 4.9 | 3.5 | 79.9 | 11.8 |
| City of Yarra | 2.8 | 4.2 | 75.0 | 18.1 |
| Frankston City Council | 2.0 | 3.0 | 73.3 | 21.8 |
| Hobsons Bay City Council | 1.5 | 4.5 | 74.6 | 19.4 |
| Hume City Council | 3.6 | 5.4 | 77.7 | 13.3 |
| Manningham Council | 1.6 | 3.2 | 88.0 | 7.2 |
| Maribyrnong City Council | 2.8 | 2.8 | 86.1 | 8.3 |
| Maroondah City Council | 2.0 | 1.0 | 79.6 | 17.3 |
| Moreland City Council | 2.1 | 4.2 | 83.3 | 10.4 |
| Mornington Peninsula Shire | 2.8 | 2.8 | 75.9 | 18.5 |
| Nillumbik Shire Council | 0.0 | 7.3 | 80.5 | 12.2 |
| Wyndham City | 2.6 | 1.6 | 83.9 | 12.0 |
| Yarra Ranges Council | 2.0 | 2.9 | 80.4 | 14.7 |
| City of Ballarat | 4.1 | 6.8 | 65.8 | 23.3 |
| City of Greater Bendigo | 3.8 | 2.5 | 68.4 | 25.3 |
| City of Greater Geelong | 2.5 | 5.0 | 67.9 | 24.5 |
| City of Latrobe | 5.3 | 5.3 | 75.4 | 14.0 |
| Greater Shepparton Council | 4.3 | 4.3 | 74.5 | 17.0 |
| Surf Coast Shire | 0.0 | 0.0 | 95.7 | 4.3 |
